# Acute Hyperkalemia and 30-Day Mortality: Increased Mortality at Slightly Elevated Plasma Potassium Levels

**DOI:** 10.64898/2026.04.10.26350589

**Authors:** Frederikke Egeberg, Hanne Nygaard, Johannes Grand, Theis Skovsgaard Itenov, Mathias Lindquist, Fredrik Folke, Helle Collatz Christensen, Jakob Lundager Forberg, Ahmad Sajadieh, Janne Petersen, Steen Bendix Haugaard, Rasmus Gregersen Mottlau

**Affiliations:** Department of Emergency Medicine, Copenhagen University Hospital – Bispebjerg and Frederiksberg, Copenhagen Denmark; Department of Cardiology, Copenhagen University Hospital – Amager and Hvidovre, Copenhagen, Denmark; Department of Cardiothoracic Anesthesiology, Copenhagen University Hospital – Rigshospitalet, Copenhagen, Denmark; Department of Cardiology, Copenhagen University Hospital Herlev and Gentofte, Denmark; Emergency Medical Services, Capital Region, Denmark; Department of Clinical Medicine, University of Copenhagen, Denmark; Prehospital Center, Region Zealand, Naestved, Denmark; Department of Emergency Medicine, Helsingborg Hospital, Helsingborg, Sweden; Department of Clinical Sciences, Lund University, Lund, Sweden; Department of Cardiology, Copenhagen University Hospital – Bispebjerg and Frederiksberg, Copenhagen, Denmark; Center for Clinical Research and Prevention, Copenhagen University Hospital – Bispebjerg and Frederiksberg, Copenhagen, Denmark; Section of Biostatistics, Department of Public Health, Faculty of Health and Medical Sciences, University of Copenhagen, Copenhagen, Denmark; Department of Endocrinology, Copenhagen University Hospital – Bispebjerg and Frederiksberg, Copenhagen, Denmark

**Author notes:** **Meetings:** Results presented at “Lassendagen”, Bispebjerg and Frederiksberg Hospital, Copenhagen, Denmark, december 5, 2025.

**Keywords:** Acute hyperkalemia, Electrolyte imbalance, Mortality, Emergency medicine

## Abstract

**Background:** Potassium is involved in multiple physiological processes in the body, and hyperkalemia is a common, potentially life-threatening condition.

**Objective:** The aim of our study was to examine the association between plasma potassium levels, and 30-day mortality in patients presenting to an emergency department with normo- or hyperkalemia.

**Design:** Retrospective Cohort study.

**Setting:** Emergency Departments in the Capital region of Denmark

**Participants:** Persons attending Emergency Departments in the Capital Region of Denmark from 2017-2021 with a plasma potassium level of at least 3.5 mM measured within 4 hours after arrival.

**Measurements:** The study was based on data from Danish National Registries and electronic patient records. We performed Kaplan-Meier survival analyses and unadjusted and adjusted cox regression analyses utilizing plasma [K+] 3.5–4.4 mM as the reference group for 30-day mortality hazard ratios (HRs).

**Results:** A total of 248,453 patients were included with a median age of 60 years (Q1;Q3 42;75), and 6,959 (2.8%) died within 30 days. Mortality was 2.2% for potassium level 3.5–4.4 mM, 6.9% for 4.5–4.9 mM, 17.1% for 5.0–5.9 mM, and 26.9% for ≥6.0 mM. Unadjusted 30-day HRs were 3.2 (95%CI: 3.0–3.4) for [K+] 4.5–4.9 mM, 8.6 (95%CI: 7.9–9.3) for [K+] 5.0–5.9 mM, and 14.7 (95%CI: 12.5–17.0) for [K+] ≥6.0 mM. Adjusted HRs were 1.4 (1.3–1.5), 2.10 (1.9–2.3), and 2.4 (2.0–2.8), respectively.

**Limitations:** Risk of residual confounding. Missing data. No access to data regarding in-hospital treatment.

**Conclusion:** Plasma potassium levels above 4.4 mM were associated with increased 30-day mortality among patients presenting to emergency departments.

**Primary funding source:** Department of Emergency Medicine, Copenhagen University hospital, Bispebjerg and Frederiksberg Hospital.

## Introduction

Hyperkalemia is one of the most common electrolyte abnormalities in patients presenting to the emergency department (ED), reported in up to 9% of the patients.(1–4) Potassium plays a regulatory role in multiple physiological mechanism in the body, including regulation of membrane potential and excitability in neurons and cardiac myocytes, pH-balance, osmotic pressure, and enzyme activities.(3) Due to the effects on the membrane potential of cardiac myocytes, hyperkalemia can cause or contribute to cardiac arrest or arrhythmia.(5) The clinical manifestations of hyperkalemia are nonspecific, including muscle weakness, paralysis, and palpitations.(6) Even patients with severe, life-threatening hyperkalemia can appear asymptomatic.(7,8) The risk of developing hyperkalemia is increased in patients with diabetes mellitus, cardiovascular disease, and chronic kidney disease.(9) Likewise, the use of certain medications, e.g., potassium supplements, spironolactone, ace-inhibitors, and angiotensin-II-receptor blockers, can cause hyperkalemia.(10)

The effect of hyperkalemia on 30-day mortality is ambiguous. Increased mortality among populations with acute hyperkalemia has been reported in those presenting with acute kidney insufficiency, infection, diabetes mellitus, use of mineralocorticoid receptor antagonist, dehydration, or bleeding.(4,11) In contrast, drug-induced hyperkalemia has been associated with lower mortality than hyperkalemia attributable to other causes.(11) Moreover, the severity of hyperkalemia has been shown to depend on the rate of potassium increase.(6) In non-acute settings, slightly elevated potassium levels have been associated with lower risk of cardiovascular arrhythmias and mortality in patients with heart failure or with an implantable cardioverter-defibrillator.(12) However, in acute settings and broader populations, evidence regarding the association between slightly elevated potassium levels and mortality remains limited. Additionally, research is challenged by the absence of universally accepted definitions of hyperkalemia.(13) The European Resuscitation Council provides treatment recommendations for plasma potassium levels ≥5.5 mM, while the clinical normal range is considered 3.5–4.4 mM.(14,15)

The aim of this study was to determine if patients presenting at emergency departments with elevated potassium levels had an increased 30-day mortality risk compared with normokalemic patients. Further, we sought to evaluate how risk differences were affected by sociodemographics, comorbidities and acute patient characteristics.

## Method

### Design and study population

This was a register-based cohort study of adult patients (≥18 years of age) arriving at an ED in the Capital Region of Denmark during a 5-year period from 2017 to 2021.(16) We obtained data from national registries and electronic patient records.

We included patient contacts of persons who were permanent residents in Denmark at time of arrival, meaning having a permanent Danish civil registration number. Additionally, we included the contacts with normo- or hyperkalemia based on results from a blood sample drawn during the first 4 hours after arrival at an ED. We utilized the first blood potassium analysis in the period if more than one was performed. The lower limit of the normal plasma potassium level was defined as 3.5 mM according to the clinical normal values.(14) To exclude pseudo-hyperkalemia, an artifact most often caused by hemolysis during phlebotomy, we excluded patient contacts with a plasma potassium of ≥6.5 mM, who subsequently had normalized potassium levels within 3 hours after the index blood test.(17) We further excluded patient contacts with prehospital cardiac arrest. Contacts with missing registration of heart rate, blood pressure, oxygen saturation, respiratory rate, body temperature, plasma sodium level, or plasma creatinine level within the first 4 hours of the ED stay were likewise excluded. Repeated eligible contacts by the same person were omitted.

### Setting

In Denmark, acute hospital care is funded by taxes and are free of charge when used. Each Danish resident is required to have a unique civil registration number utilized by all national authorities, which enables individual-level linkage across the different national registries containing various types of personal information.(18,19) Since 2007, the Danish healthcare services have been administered separately within each of the five regions of Denmark. The Capital Region of Denmark has approximately 1,700,000 inhabitants. The patients are commonly referred by their general practitioner, or through telephonic contact with the regional medical helpline, or the national emergency helpline.(18)

### Outcome

The outcome of the study was 30-day all-cause mortality from the ED arrival date.

### Acute patient characteristics

The first potassium measurement for each patient contact was used to stratify the patients into the reference group of normokalaemia (3.5–4.4 mM), and 3 exposure groups – slightly elevated potassium level (4.5–4.9 mM), mild hyperkalemia (5.0–5.9 mM), and moderate to severe hyperkalemia (≥6.0 mM).

The acute patient characteristics included information obtained during the first 4 hours after arrival, regarding heart rate, systolic blood pressure, body temperature, respiratory rate, plasma sodium level, and plasma creatinine level. The mentioned data was grouped based on normal ranges, except plasma sodium and creatinine level. Additionally, acute characteristics included information regarding the type of transport to the ED. The way of arrival was grouped into “Ambulance priority A or helicopter”, “Ambulance priority B or C”, “Arrival without help or by seated transport”, and “Unspecified”. Further, signs of infection were included. An arrival C-reactive protein ≥10 mg/L, and/or blood leukocyte count ≥10 mia/L, and/or a registered body temperature ≥38°C were defined as signs of infection. Acute characteristics further included information regarding the patients do-not-resuscitate order status at ED arrival date.

### Sociodemographics and comorbidities

Sociodemographic characteristics included age, sex, cohabitation, and educational level. Cohabitation status at arrival specified whether the patient was living alone or in a shared household. Information regarding the patient’s highest level of education was grouped by the International Standard Classification of Education (ISCED).(20) For multimorbidity evaluation, we assessed diagnosis codes from 10 years before ED arrival, using the Measuring MultiMorbidity index (M3-index).(21) We evaluated selected comorbidities from the M3-index: chronic kidney disease (CKD), myocardial infarction, cardiac arrhythmias, diabetes mellitus (DM), and major psychiatric diagnoses (Supplementary S3: M3 diagnosis codes). We identified if the patient was previously known to have hyperkalemia, defined as a registered plasma potassium level ≥5.5 mM, in the period 1 year prior to ED arrival until 24 hours before arrival. We evaluated information regarding redeemed medication prescriptions within 6 months prior to ED arrival by Anatomical Therapeutic Chemical Classification System (ATC) codes for the drug groups: NSAID, renin-angiotensin-aldosterone-system (RAAS) acting agents, beta blockers or digoxin, potassium supplement, and glucocorticoids. Additionally, we assessed information regarding redeemed prescriptions of antibiotics within 30 days prior to arrival (Supplementary S4: ATC codes).

### Data sources

This study utilized data from national registries, including the Civil Registration System (CPR), the Danish National Patient Register (DNPR) (22), the Danish National Prescription Register (23), Register of Laboratory Results for Research (24), and the Danish Population Education Register(25). We included data from electronic patient records: Computer Assisted Dispatch (information regarding prehospital phone calls), Prehospital Patient Journal (used by prehospital medical services), and Sundhedsplatformen delivered by Epic Systems (the electronic patient record used by hospitals in the Capital Region of Denmark) (18). The obtained information from each source was presented in Supplementary table S1.

### Statistical analysis

Age, p-sodium and p-creatinine were presented by median and quartiles(Q1;Q3), while the remaining acute and baseline characteristics, were presented by frequency and percentages. The unadjusted absolute 30-day survival probability in each potassium level group was visualized using a Kaplan-Meier survival function. We did a locally estimated scatterplot smoothing of the Kaplan-Meier plot, due to protection of microdata rules from Statistics Denmark. Unadjusted and adjusted cox regression analyses were performed, determining hazard ratios (HRs) with 95% confidence intervals (CIs) for 30-day mortality in each potassium group, using normokalemia (3.5–4.4 mM) as the reference group. We did a directed acyclic graph (DAG) to determine relevant confounding co-variables (figure not shown), adjusting for the above-mentioned characteristics, including socio-demographics, comorbidities, and acute characteristics. Further, we performed time-stratified cox regression analyses, determining the unadjusted and adjusted mortality risk for each potassium group, within different time stratifications (days 0–1, 2–7, and 8–30).

## Results

### Population

This study included 248,453 patients (Figure 1). Compared with the normokalemic group, patients in the hyperkalemic groups were older, more frequently males, had a higher M3-index, and more often had CKD, myocardial infarction, arrhythmia, and DM (Table 1). The hyperkalemic patients more frequently redeemed prescriptions for RAAS acting agents, beta-blockers or digoxin, potassium supplements, glucocorticoids, and antibiotics. They were more often living by themselves, had a lower level of education, and had a history of hyperkalemia within 1 year prior to ED arrival. At ED arrival, the hyperkalemic group more often had a heart rate out of the normal range, low systolic blood pressure, low oxygen saturation, high respiratory rate, low body temperature, and had a more acute way of arrival. Additionally, they more frequently had a do-not-resuscitate order, low plasma sodium levels, elevated plasma creatinine levels, and signs of infection (Table 2).

**Table 1.** Characteristics of normo- and hyperkalemic ([K+] ≥3.5 mM) persons attending an emergency department in the Capital Region of Denmark from 2017 to 2021.

| Characteristic | Group | Total | [K<sup>+</sup>] 3.5–4.4 mM | [K<sup>+</sup>] 4.5–4.9 mM | [K<sup>+</sup>] 5.0–5.9 mM | [K<sup>+</sup>] ≥6.0 mM |
| --- | --- | --- | --- | --- | --- | --- |
| Total |  | 248,453 | 227,312<br>(91.5%) | 16,718<br>(6.7%) | 3,802<br>(1.5%) | 621<br>(0.2%) |
| Sex | Female | 126,622<br>(51.0%) | 118,025<br>(51.9%) | 6,774<br>(40.5%) | 1,556<br>(40.9%) | 267<br>(43.0%) |
| Age |  | 60.0<br>(42.0;75.0) | 58.0<br>(41.0;74.0) | 72.0<br>(58.0;82.0) | 76.0<br>(67.0;84.0) | 75.0<br>(67.0;85.0) |
| M3-index | 0 to <1 | 198,155<br>(79.8%) | 185,093<br>(81.4%) | 10,796<br>(64.6%) | 1,983<br>(52.2%) | 283<br>(45.6%) |
|  | 1 to <2 | 35,315<br>(14.2%) | 30,030<br>(13.2%) | 3,920<br>(23.4%) | 1,165<br>(30.6%) | 200<br>(32.2%) |
|  | 2+ | 14,983<br>(6.0%) | 12,189<br>(5.4%) | 2,002<br>(12.0%) | 654<br>(17.2%) | 138<br>(22.2%) |
| Comorbidities | CKD* | 7,057<br>(2.8%) | 5,126<br>(2.3%) | 1,249<br>(7.5%) | 567<br>(14.9%) | 115<br>(18.5%) |
|  | Myocardial infarction | 10,089<br>(4.1%) | 8,481<br>(3.7%) | 1,189<br>(7.1%) | 363<br>(9.5%) | 56<br>(9.0%) |
|  | Arrhythmia | 32,038<br>(12.9%) | 26,979<br>(11.9%) | 3,779<br>(22.6%) | 1,099<br>(28.9%) | 181<br>(29.1%) |
|  | DM* | 21,562<br>(8.7%) | 17,051<br>(7.5%) | 3,164<br>(18.9%) | 1,141<br>(30.0%) | 206<br>(33.2%) |
|  | Major psychiatric diagnosis | 17,554<br>(7.1%) | 16,007<br>(7.0%) | 1,205<br>(7.2%) | 294<br>(7.7%) | 48<br>(7.7%) |
| Previous hyperkalemic |  | 9,509<br>(3.8%) | 5,760<br>(2.5%) | 2,286<br>(13.7%) | 1,233<br>(32.4%) | 230<br>(37.0%) |
| Redeemed drug prescriptions | Antibiotics | 34,037<br>(13.7%) | 30,149<br>(13.3%) | 2,910<br>(17.4%) | 835<br>(22.0%) | 143<br>(23.0%) |
|  | NSAID<sup>†</sup> | 33,461<br>(13.5%) | 30,697<br>(13.5%) | 2,263<br>(13.5%) | 431<br>(11.3%) | 70<br>(11.3%) |
|  | RAAS<sup>‡</sup> acting agent | 60,729<br>(24.4%) | 51,428<br>(22.6%) | 6,836<br>(40.9%) | 2,081<br>(54.7%) | 384<br>(61.8%) |
|  | Betablocker or digoxin | 38,440<br>(15.5%) | 31,689<br>(13.9%) | 4,938<br>(29.5%) | 1,536<br>(40.4%) | 277<br>(44.6%) |
|  | Potassium supplement | 18,715<br>(7.5%) | 15,411<br>(6.8%) | 2,282<br>(13.6%) | 811<br>(21.3%) | 211<br>(34.0%) |
|  | Glucocorticoid | 11,948<br>(4.8%) | 10,351<br>(4.6%) | 1,196<br>(7.2%) | 347<br>(9.1%) | 54<br>(8.7%) |
| Cohabitation | Living alone | 90,467<br>(36.4%) | 80,652<br>(35.5%) | 7,480<br>(44.7%) | 1,991<br>(52.4%) | 344<br>(55.4%) |
| Highest level of education | Missing or ISCED 0–2<sup>‡</sup> | 76,288<br>(30.7%) | 68,640<br>(30.2%) | 5,923<br>(35.4%) | 1,472<br>(38.7%) | 253<br>(40.7%) |
|  | ISCED 3–4<sup>‡</sup> | 96,998<br>(39.0%) | 88,558<br>(39.0%) | 6,636<br>(39.7%) | 1,563<br>(41.1%) | 241<br>(38.8%) |
|  | ISCED 5–8<sup>‡</sup> | 75,167<br>(30.3%) | 70,114<br>(30.8%) | 4,159<br>(24.9%) | 767<br>(20.2%) | 127<br>(20.5%) |

\*CKD, Chronic Kidney Disease; DM, Diabetes Mellitus.
<sup>†</sup>NSAID, Nonsteroidal anti-inflammatory drug; RAAS acting agent, renin-angiotensin-aldosterone system acting agent.
<sup>‡</sup>ISCED, International Standard Classification of Education; ISCED 0–2, less than primary, primary or lower secondary; ISCED 3–4, Upper secondary or post-secondary, non-tertiary; ISCED 5–8, Short-cycle tertiary or above.

**Table 2.** Acute characteristics of normo- and hyperkalemic ([K+] ≥3.5 mM) persons attending an emergency department in the Capital Region of Denmark from 2017 to 2021.

| Characteristic | Group | Total | [K+] 3.5–4.4 mM | [K+] 4.5–4.9 mM | [K+] 5.0–5.9 mM | [K+] 6.0 mM |
| --- | --- | --- | --- | --- | --- | --- |
| **Total** |  | 248,453 | 227,312 (91.5%) | 16,718 (6.7%) | 3,802 (1.5%) | 621 (0.2%) |
| **Heart rate** | **<60 beats/min** | 18,204 (7.3%) | 16,251 (7.1%) | 1,471 (8.8%) | 366 (9.6%) | 116 (18.7%) |
|  | **60–100 beats/min** | 190,076 (76.5%) | 174,863 (76.9%) | 12,218 (73.1%) | 2,615 (68.8%) | 380 (61.2%) |
|  | **>100 beats/min** | 40,173 (16.2%) | 36,198 (15.9%) | 3,029 (18.1%) | 821 (21.6%) | 125 (20.1%) |
| **Systolic Blood Pressure** | **<100 mmHg** | 5,869 (2.4%) | 4,754 (2.1%) | 638 (3.8%) | 360 (9.5%) | 117 (18.8%) |
|  | **100–140 mmHg** | 133,525 (53.7%) | 122,506 (53.9%) | 8,658 (51.8%) | 2,036 (53.6%) | 325 (52.3%) |
|  | **>140 mmHg** | 109,059 (43.9%) | 100,052 (44.0%) | 7,422 (44.4%) | 1,406 (37.0%) | 179 (28.8%) |
| **Oxygen saturation** | **<88%** | 1,668 (0.7%) | 1,282 (0.6%) | 255 (1.5%) | 98 (2.6%) | 33 (5.3%) |
|  | **88–95%** | 19,428 (7.8%) | 16,759 (7.4%) | 1,990 (11.9%) | 580 (15.3%) | 99 (15.9%) |
|  | **>95%** | 227,357 (91.5%) | 209,271 (92.1%) | 14,473 (86.6%) | 3,124 (82.2%) | 489 (78.7%) |
| **Body temperature** | **<35.5°C** | 488 (0.2%) | 359 (0.2%) | 59 (0.4%) | 51 (1.4%) | 19 (3.1%) |
|  | **35.5–37.4°C** | 210,504 (86.4%) | 191,974 (86.2%) | 14,651 (89.1%) | 3,327 (88.9%) | 552 (89.8%) |
|  | **37.5–38.5°C** | 17,391 (7.1%) | 16,168 (7.3%) | 987 (6.0%) | 215 (5.7%) | 21 (3.4%) |
|  | **>38.5°C** | 15,212 (6.2%) | 14,284 (6.4%) | 754 (4.6%) | 151 (4.0%) | 23 (3.7%) |
| **Respiratory rate** | **≤16/min** | 129,901 (52.3%) | 120,326 (52.9%) | 7,928 (47.4%) | 1,461 (38.4%) | 186 (30.0%) |
|  | **>16/min** | 118,552 (47.7%) | 106,986 (47.1%) | 8,790 (52.6%) | 2,341 (61.6%) | 435 (70.0%) |
| **Type of Arrival** | **Priority A** | 24,855 (10.0%) | 22,655 (10.0%) | 1,699 (10.2%) | 411 (10.8%) | 90 (14.5%) |
|  | **Priority B or C** | 58,278 (23.5%) | 52,420 (23.1%) | 4,401 (26.3%) | 1,247 (32.8%) | 210 (33.8%) |
|  | **Without help or seated transport** | 128,335 (51.7%) | 118,474 (52.1%) | 8,123 (48.6%) | 1,524 (40.1%) | 214 (34.5%) |
|  | **Unspecified** | 36,985 (14.9%) | 33,763 (14.9%) | 2,495 (14.9%) | 620 (16.3%) | 107 (17.2%) |
| **Do-not-resuscitate order** |  | 2,024 (0.8%) | 1,568 (0.7%) | 283 (1.7%) | 149 (3.9%) | 24 (3.9%) |
| **p-[Sodium]** |  | 139 (137; 141) | 139 (137; 141) | 138 (135; 140) | 137 (133; 140) | 134 (130; 138) |
| **p-[Creatinine]** |  | 74.0 (62.0; 89.0) | 73.0 (62.0; 87.0) | 89.0 (72.0; 119) | 128 (89.0; 200) | 226 (153; 434) |
| **Sign of Infection** |  | 123,994 (49.9%) | 111,030 (48.8%) | 9,746 (58.3%) | 2,708 (71.2%) | 510 (82.1%) |

**Figure 1.**
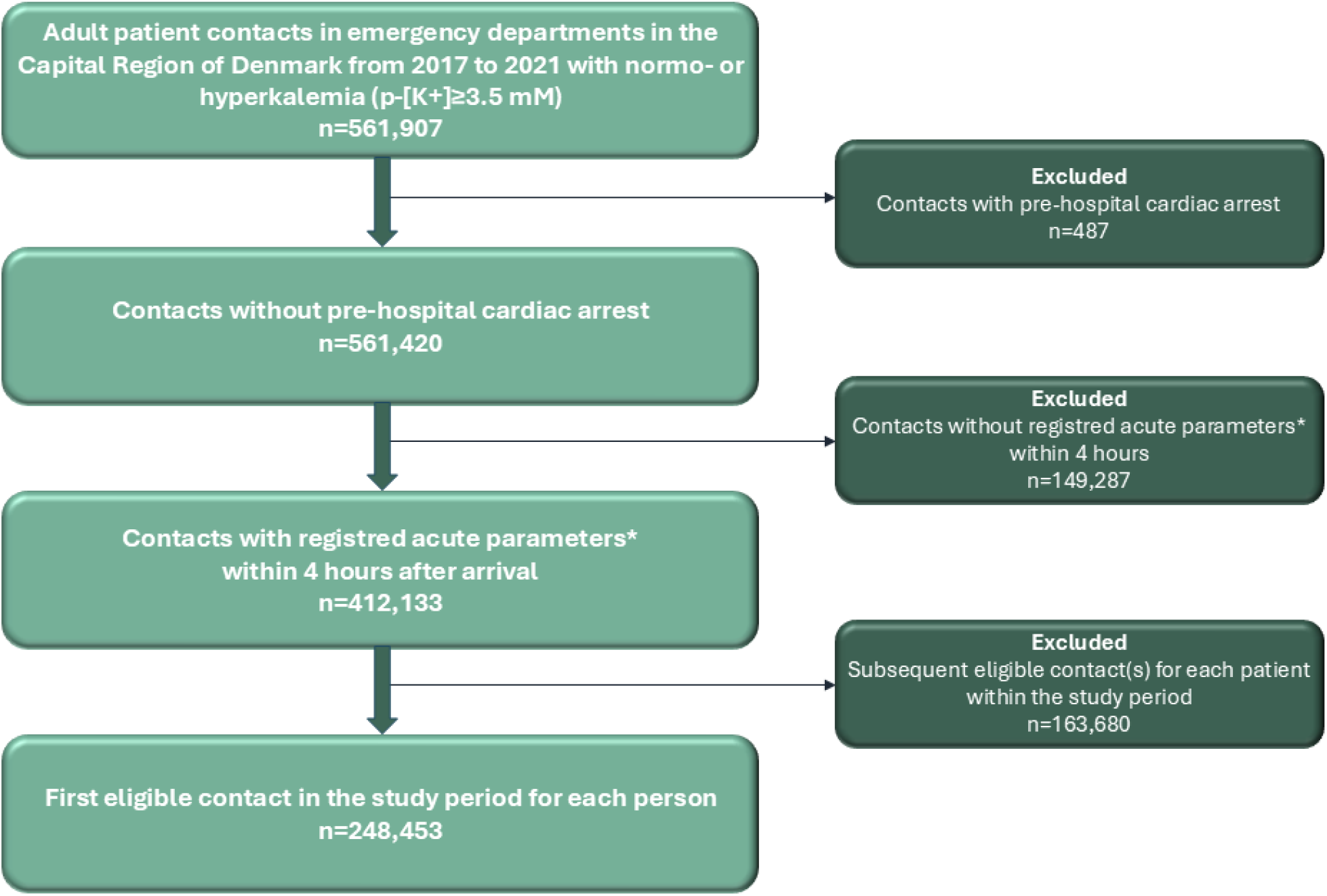
Patient population flowchart (inclusions and exclusions) *Acute parameters include heart rate, blood pressure, oxygen saturation, respiratory rate, body temperature, plasma sodium level, and plasma creatinine level.

### Mortality risk

In our population 6,959 (2.8%) patient contacts lead to mortality within 30-days. Of those, 1,964 (28.2%) patients had hyperkalemia at ED arrival (Table 3). The unadjusted 30-day mortality risks were presented in a statistically smoothened Kaplan-Meier plot in Figure 2. The plot demonstrated an increased 30-day mortality risk with increasing plasma potassium levels, with most deaths occurring within 7 days. Mortality estimates were 2.2% (95%CI: 2.1%–2.3%) for [K+] 3.5–4.4mM, 6.9% (95%CI: 6.5%–7.3%) for [K+] 4.5–4.9 mM, 17.1% (95%CI: 16.0%–18.4%) for [K+] 5.0–5.9 mM, and 26.9% (95%CI: 23.6 %–30.6%) for [K+] ≥6.0 mM.

**Table 3.** Unadjusted and adjusted* hazard ratios (HRs) for mortality within different time intervals following emergency department arrival of normo- and hyperkalemic ([K+] ≥3.5 mM) persons attending an emergency department in the Capital Region of Denmark from 2017 to 2021.The HRs are presented for different potassium level groups using normokalemia (3.5–4.4 mM) as the reference with a 95% confidence interval (CI).

|  | [K+] 4.5–4.9 mM | [K+] 5.0–5.9 mM | [K+] ≥6.0 mM |
| --- | --- | --- | --- |
| **Unadjusted 30-day mortality**<br>**HR (95%CI)** | 3.20 (3.00–3.41) | 8.55 (7.87–9.27) | 14.66 (12.52–17.04) |
| **Adjusted\* 30-day mortality**<br>**HR (95%CI)** | 1.40 (1.31–1.50) | 2.10 (1.91–2.29) | 2.36 (1.97–2.80) |
| **Unadjusted mortality risk within 0–1 day**<br>**HR (95%CI)** | 3.26 (2.62–4.02) | 12.65 (10.03–15.77) | 36.53 (26.23–49.49) |
| **Adjusted\* mortality risk within 0–1 day**<br>**HR (95%CI)** | 1.46 (1.16–1.81) | 2.72 (2.10–3.48) | 3.94 (2.62–5.79) |
| **Unadjusted mortality risk within 2–7 days, if alive at the beginning of day 2**<br>**HR (95%CI)** | 3.43 (3.04–3.86) | 10.52 (9.11–12.08) | 15.89 (11.90–20.73) |
| **Adjusted\* mortality risk within 2–7 days, if alive at the beginning of day 2**<br>**HR (95%CI)** | 1.48 (1.31–1.68) | 2.56 (2.19–2.99) | 2.57 (1.86–3.48) |
| **Unadjusted mortality risk within 8–30 days if alive at the beginning of day 8**<br>**HR (95%CI)** | 1.03 (1.01–1.05) | 1.09 (1.05–1.12) | 1.14 (1.05–1.24) |
| **Adjusted\* mortality risk within 8–30 days if alive at the beginning of day 8**<br>**HR (95%CI)** | 1.01 (1.00–1.03) | 1.04 (1.01–1.08) | 1.07 (0.97–1.17) |

\*Adjusted for age, sex, M3-index, Chronic Kidney Disease, Myocardial Infarction, Cardiac Arrhythmias, Diabetes Mellitus and Major Psychiatric Diagnoses, previously hyperkalemic, cohabitation status, educational level, redeemed drug prescriptions of NSAID, RAAS acting agents, Beta blocker or digoxin, potassium supplement, and glucocorticoid prescriptions redeemed within 6 months prior to emergency department arrival, and antibiotic within 30 days prior to emergency department arrival. Further, heart rate, systolic blood pressure, body temperature, oxygen saturation, respiratory rate, plasma sodium level, plasma creatinine level, way of arrival to the ED, sign of infection, and do-not-resuscitate order at arrival.

**Figure 2.**
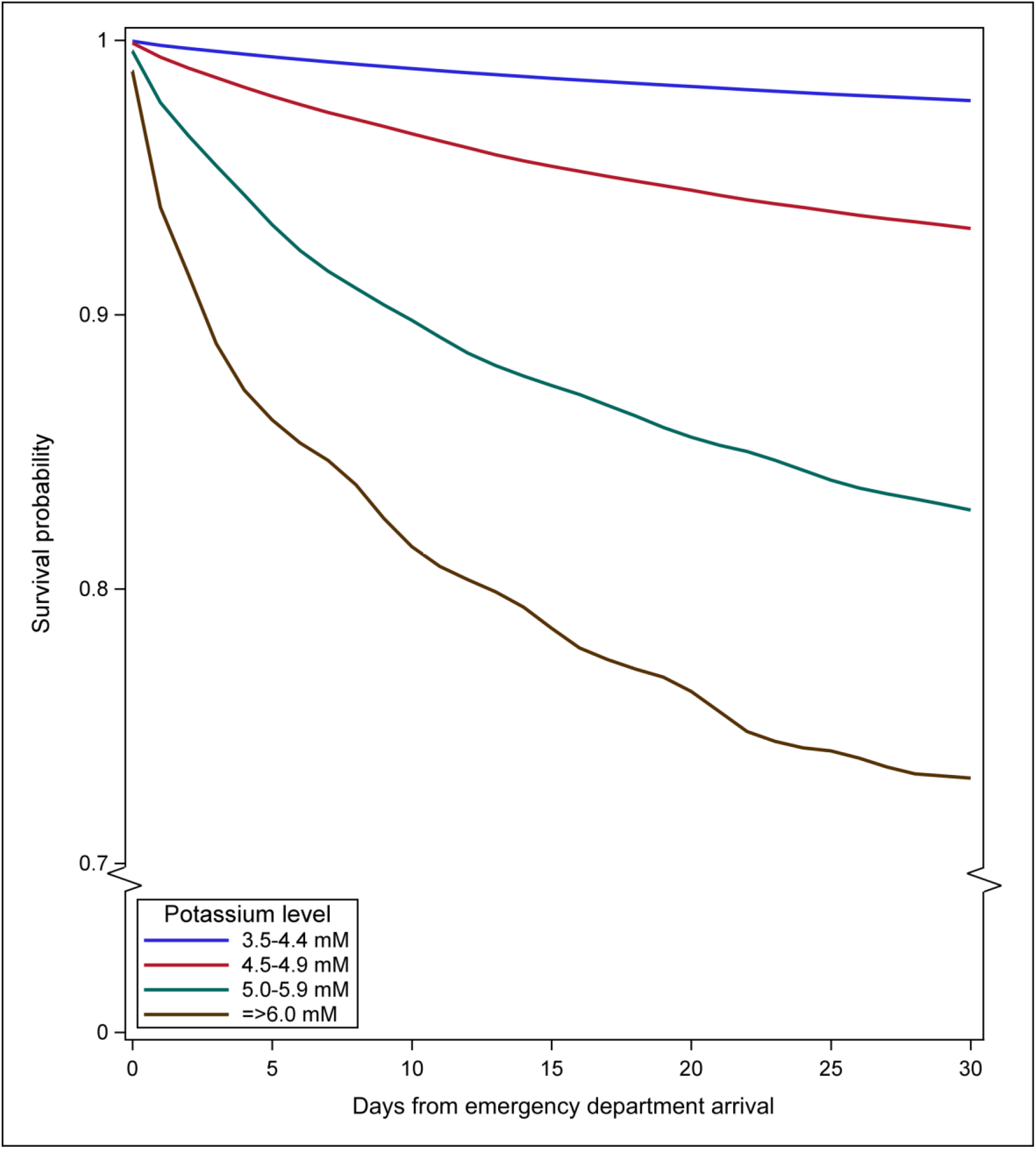
Statistically smoothened* Kaplan-Meier plot illustrating the association between different plasma potassium levels and absolute 30-day survival. The plot presents the data with bracketed y-axis. *A locally estimated scatterplot smoothing of the Kaplan-Meier plot was made due to protection of microdata rules from Statistics Denmark

All hyperkalemic groups had an increased 30-day mortality risk, compared to the normokalemic group. The relative difference in risk corresponds to unadjusted HRs at 3.2 (95%CI: 3.0-3.4) for [K+] 4.5–4.9 mM, 8.6 (95%CI: 7.9–9.3) for [K+] 5.0–5.9 mM, and 14.7 (95%CI: 12.5–17.0) for [K+] ≥6.0 mM, using normokalemia as reference group. After adjustments, the HRs diminished, but the mortality risk remained significantly increased in all hyperkalemic groups. Adjusted HRs were 1.4 (95%CI: 1.3–1.5) for [K+] 4.5–4.9 mM, 2.1 (95%CI: 1.9–2.3) for [K+] 5.0–5.9 mM, and 2.4 (2.0–2.8) for [K+] ≥6.0 mM (Table 3).

We observed non-proportional hazards within 30 days with an increased risk of mortality the first 7 days. Hazard ratio for the p-[K+] 4.5–4.9 mM group was 1.5 (95%CI 1.2–1.8) for mortality within 0– 1 day after ED arrival, 1.5 (95%CI: 1.3–1.7) for mortality within 2–7 days, and 1.0 (95%CI: 1.0–1.0) within 8–30 days. Likewise, in the other hyperkalemic groups, an increased mortality risk within 0– 1 day and 2–7 days was identified. Unadjusted and adjusted HRs to the different time stratifications were presented in Table 3.

## Discussion

### Major findings

This study demonstrated an association between plasma potassium levels of 4.5 mM or more and increased 30-day mortality among persons presenting at an ED. The association remained significant after adjustments for several available confounders, although the HRs were attenuated. Time-stratified analyses revealed that the mortality risk among hyperkalemic patients with hyperkalemia was markedly higher than in normokalemic patients during the first seven days following ED arrival.

Previous studies have reported comparable associations to those observed in the present study. One study included all adults in Manitoba, Canada, with de novo hyperkalemia between 1 January 2007 and 31 December 2016. They assessed the relative 30-day mortality risk in 88,541 patients with [K+] ≥5.5 mM compared with a matched normokalemic reference group with [K+] <5.0 mM. The study reported an unadjusted 30-day mortality HR of 3.6 and an adjusted HR of 1.3, adjusting for age, sex, comorbidities, and medication use.(26) In our study, adjustment for confounders reduced HRs from 3.2 to 1.4, 8.6 to 2.1, and 14.7 to 2.4, respectively (Table 3).

Most previous studies on hyperkalemia defined the condition using higher plasma potassium thresholds than in the present study. The Canadian study defined hyperkalemia as [K+] ≥5.0 mM, while a 2017 American ED study of in-hospital mortality among hyperkalemic patients used [K+] 3.5–4.9 mM as the normokalemic reference group.(26–29) In our study, persons with slightly elevated potassium levels of 4.5–4.9 mM had a 40% higher adjusted risk of mortality compared with the reference group ([K+] 3.5–4.4 mM). These findings propose that an association between higher potassium levels and short-term mortality among patients presenting at an ED is evident even before potassium levels exceeds 4.9 mM. An association between plasma potassium levels of 4.5– 5.0 mM and mortality has been reported in various disease-specific populations, e.g., patients with acute heart failure, acute myocardial infarction, and cardiovascular diseases.(4,30,31) Studies including broader, unselected emergency department populations is less frequent. Among the limited evidence is a 2017 study from a hospital in South Korea, which included 17,777 hospitalized patients. The study reported an increased 30-day mortality risk in patients with plasma potassium levels of ≥4.6 mM.(32) After adjustment for age, sex, comorbidities, medication use, and selected laboratory results, the reported 30-day mortality HRs were 1.6 for [K+] 4.6–5.0 mM, 2.2 for [K+] 5.1– 5.5 mM, and 4.1 for [K+]>5.5 mM, utilizing [K+] 3.6–4.0 as reference group. The South Korean study demonstrated an association between 30-day mortality risk and potassium levels on 4.6 mM or more, aligning with our findings in persons presenting at Danish EDs.

### Strengths and limitations

This study was based on data from Danish national registers and electronic patient records. The Danish national registers are considered to have a high validity and completeness (22,25,33). The study design allowed inclusion of a large population of 248,453 patients, providing strong statistical power. The inclusion criteria (ED contacts with normo- or hyperkalemia), exposure (high plasma potassium level), and outcome (30-day mortality) were all independent of clinical registration, resulting in a low risk of selection bias. Due to the nature of the study, the data were collected prior to the initiation of the study, minimizing the risk of bias during data collection. During population selection, a total of 149,287 patient contacts were excluded because of missing data within the first 4 hours after ED arrival. The missing data are presented in Supplementary S6. The missing data were evenly distributed across hospitals and years, and no systematic pattern was detected.

The blood samples used in this study were collected by various clinicians for different clinical indications, and analyses were performed using various laboratory equipment, which may have introduced minor variability in the measurements. Some blood analyses, including potassium measurements, were performed using point-of-care equipment, and we did not have access to all point-of-care data. If hyperkalemia was detected on a point-of-care analysis, standard practice was to perform a confirmatory laboratory analysis. All such analyses were available in the Register of Laboratory Results for Research, which was utilized in this study. Another source of uncertainty regarding plasma potassium levels was the risk of pseudo-hyperkalemia – an artifact causing falsely high plasma potassium levels, often resulting from hemolysis during phlebotomy.(17) We strove to account for this phenomenon by excluding 52 ED visits with a plasma potassium level of more than 6.5 mM at arrival, and a subsequent measurement under 4.5 mM within 3 hours. Nevertheless, it is possible that not all pseudo-hyperkalemic patients were excluded, and there is a risk that we unintentionally excluded patients who received fast treatment in the ED, resulting in rapid potassium normalization.(34) We did not have access to data on in-hospital treatment, which could have improved the identification of pseudo-hyperkalemic patients by cross-checking the initiation of hyperkalemic treatment. However, the number of pseudo-hyperkalemic patients is expected to be low and is therefore unlikely to affect the study conclusions. Regarding drug consumption prior to ED arrival, we had access to data on whether the patient redeemed a prescription at a pharmacy in Denmark, but no information on compliance. In most cases, redeemed prescriptions reflect consumption, although this could not be confirmed.

We did not find proportional hazards through 30 days; thus, we assessed the differentiated effect by time-stratified analyses, which showed a higher mortality among the hyperkalemic patients within the first 7 days after arrival. A high mortality in the beginning of the period is expected in studies like ours involving acute conditions that require immediate intervention. Consequently, the main results in our study represent an average across the 30-day outcome period.

Finally, as with other observational studies, residual confounding could not be fully accounted for, and we did not know whether the causes of death were hyperkalemia, or another condition. However, we adjusted for several relevant covariables, determined with the use of a DAG.

### Perspectives and clinical implications

In our study population, 30-day mortality risk was significantly increased at all levels of hyperkalemia, including slightly increased plasma potassium levels, i.e., 4.5–4.9 mM. The hyperkalemia guideline by the European Resuscitation Council recommends considering monitoring and treatment at potassium levels on 5.5 mM or more (15). Our study presented an increased risk of mortality at potassium levels lower than 5.5 mM, suggesting that preventive action may be relevant at lower potassium levels. This will need further testing in clinical studies. It is possible that available treatment options are ineffective when potassium is below 5.5 mM, or that slightly elevated hyperkalemia more often reflects an underlying condition contributing to increased mortality. The evidence is scarce, and potentially reducing this excess mortality should be a subject for future studies. Identifying the causes of hyperkalemia and death within each potassium group could improve understanding the impact of different potassium levels and potentially insight on strategies for reducing the mortality risk. Furthermore, as hyperkalemia predisposes to arrhythmias, incorporating electrocardiogram findings in future studies would be valuable (17).

## Conclusion

The study demonstrated an association between plasma potassium levels above 4.4 mM and increased 30-day mortality. The association diminished but was robust to confounder adjustment at all levels of hyperkalemia. The increased mortality risk was most pronounced within the first 7 days after ED arrival.

## Data Availability

Due to the rules of protection of individual's data from Statistics Denmark, it is not possible to share the data in any raw or anonymized form. Danish research institutions can obtain permission to access the data on equal term.

## Abbreviations

ED: Emergency Department
DAG: Directed Acyclic Graph
ISCED: International Standard Classification of Education
CKD: Chronic Kidney Disease
DM: Diabetes Mellitus
ATC: Anatomical Therapeutic Chemical
RAAS: Renin-Angiotensin-Aldosterone-system
CPR: Civil Personal Registration
DNPR: Danish National Patient Register
CI: Confidence Interval
HR: Hazard Ratio.

## Ethics Approval

Ethical approval is not required for registry-based studies in Denmark. The project was approved by Statistics Denmark and the Data Protection Agency. Data management was performed in SAS Enterprise on the servers of Statistics Denmark.

