## Supplementary Material S1-S5 for "Acute Hyperkalemia and 30-Day Mortality: Increased Mortality at Slightly Elevated Plasma Potassium Levels"

### An Observational Study

**Authors**: Frederikke Egeberg[1], Hanne Nygaard[1], Johannes Grand[2], Theis Skovsgaard Itenov[3], Mathias Lindquist[1], Fredrik Folke[4,5,6], Helle Collatz Christensen[6,7], Jakob Lundager Forberg[8,9], Ahmad Sajadieh[6,10], Janne Petersen[11,12], Steen Bendix Haugaard[6,13], Rasmus Gregersen Mottlau[1,11].

**Affiliations**

[1] Department of Emergency Medicine, Copenhagen University Hospital – Bispebjerg and Frederiksberg, Copenhagen Denmark

[2] Department of Cardiology, Copenhagen University Hospital – Amager and Hvidovre, Copenhagen, Denmark

[3] Department of Cardiothoracic Anesthesiology, Copenhagen University Hospital – Rigshospitalet, Copenhagen, Denmark.

[4] Department of Cardiology, Copenhagen University Hospital Herlev and Gentofte, Denmark

[5] Emergency Medical Services, Capital Region, Denmark.

[6] Department of Clinical Medicine, University of Copenhagen, Denmark

[7] Prehospital Center, Region Zealand, Naestved, Denmark.

[8] Department of Emergency Medicine, Helsingborg Hospital, Helsingborg, Sweden.

[9] Department of Clinical Sciences, Lund University, Lund, Sweden.

[10] Department of Cardiology, Copenhagen University Hospital – Bispebjerg and Frederiksberg, Copenhagen, Denmark.

[11] Center for Clinical Research and Prevention, Copenhagen University Hospital – Bispebjerg and Frederiksberg, Copenhagen, Denmark

[12] Section of Biostatistics, Department of Public Health, Faculty of Health and Medical Sciences, University of Copenhagen, Copenhagen, Denmark

[13] Department of Endocrinology, Copenhagen University Hospital – Bispebjerg and Frederiksberg, Copenhagen, Denmark

**Table S1** Data included in the study and the source of origin (register or electronic patient record)

| Register | Information |
|---|---|
| The Civil Registration System (CPR) | Sex, age, cohabitation status, and date of death. |
| The Danish National Patient Register (DNPR) | ED visits arrival time, and comorbidities. |
| Danish National Prescription Register | Redeemed drug prescriptions |
| Register of Laboratory Results for Research (RLRR) | Plasma potassium level, plasma sodium level, plasma creatinine level, C-reactive protein level, and blood leukocyte count. |
| Danish Population Education Register | Highest level of education |
| **Electronic patient record** | **Information** |
| Prehospital Patient Journal (PPJ) | Prehospital cardiac arrest |
| Sundhedsplatformen delivered by Epic Systems (SP) | Heart rate, systolic blood pressure, oxygen saturation, body temperature, respiratory rate, and do-not-resuscitate order. Way of arrival (missing data were covered first by PPJ, second by Computer Assisted Dispatch (CAD)) |

**Table S2** Hospital departments included in the study and the corresponding SHAK* code.

| SHAK*-code | Hospital department | Number of contacts |
|---|---|---|
| 1301288 | Rigshospitalet, Traumecenter og akutmodtagelse | 3,334 (1.3%) |
| 130185X | Rigshospitalet, Akutklinikken Glostrup | 15,939 (6.4%) |
| 1309470 | Bispebjerg og Frederiksberg hospital, Medicinsk modtageafdeling | 6,372 (2.6%) |
| 1309478 | Bispebjerg og Frederiksberg hospital, Skadeafdeling | 25,376 (10.2%) |
| 130947A | Bispebjerg og Frederiksberg hospital, Akutklinik FRH | 255 (0.1%) |
| 130947D | Bispebjerg og Frederiksberg hospital, Akutmodtagelse, medicinsk deldøgnsafdeling | 915 (0.4%) |
| 1309695 | Bispebjerg og Frederiksberg hospital, Medicinsk Modtageafdeling FRH | 2,827 (1.1%) |
| 1309698 | Bispebjerg og Frederiksberg hospital, Akutklinik FRH | 1,584 (0.6%) |
| 1330190 | Amager og Hvidovre hospital, Akut modtageafdeling Hvidovre hospital | 44 (0.02%) |
| 1330324 | Amager og Hvidovre hospital, Akutklinik observation AMH | 11,138 (4.5%) |
| 133032A | Amager og Hvidovre hospital, Akutklinik AMH | 13,419 (5.4%) |
| 1330628 | Amager og Hvidovre hospital, Skadestuen Hvidovre hospital | 14,535 (5.9%) |
| 1516360 | Herlev og Gentofte hospital, Akutmodtagelse A sengeafd. | 8 (<0.01%) |
| 1516367 | Herlev og Gentofte hospital, Overafdeling Akutmodtagelsen A, Akutklinik GE | 536 (0.2%) |
| 1516368 | Herlev og Gentofte hospital, Skadestue | 52,051 (21.0%) |
| 1516438 | Herlev og Gentofte hospital, Medicinsk overafdeling C, Akutklinik GE | 26,778 (10.8%) |
| 2000170 | Hospitalerne i Nordsjælland, HI Akutafdeling senge | 14 (<0.01%) |
| 2000171 | Hospitalerne i Nordsjælland, FS akutklinik, senge | 3 (<0.01%) |
| 2000177 | Hospitalerne i Nordsjælland, SH akutklinik, skadestue | 450 (0.2%) |
| 2000178 | Hospitalerne i Nordsjælland, HI Akutafdeling Skadestue | 57,626 (23.2%) |
| 2000179 | Hospitalerne i Nordsjælland, FS Akutklinik, Skadestue | 4,848 (2.0%) |
| 4001108 | Bornholms Hospital, Akutmodtagelsen | 10,401 (4.2%) |

*SHAK: Hospital Department Classification

**Table S3** M3-index* disease groups and ICD10† diagnosis codes.

| Disease group (ICD10† group code) | ICD10† diagnosis codes |
|---|---|
| **Chronic renal (ICD015)** | DI129, DI139, DQ60-DQ609, DQ611-DQ613, DN032, DN033, DN034, DN035, DN036, DN037, DN038, DN039, DN042, DN043, DN044, DN045, DN046, DN047, DN048, DN049, DN052, DN053, DN054, DN055, DN056, DN057, DN058, DN059, DN11-DN119, DN18-DN1899, DN19-DN199, DN250, DN258, DN259, DI120, DI131, DZ490-DZ499, DZ940, DZ992 |
| **Myocardial Infarction (ICD001)** | DI21-DI219, DI22-DI229, DI23-DI239, DI241, DI252 |
| **Cardiac arrhythmia (ICD030)** | DI441, DI442, DI443, DI456, DI459, DI47-DI479, DI48-DI489, DI49-DI499, DT821, DZ450, DZ950 |
| **Diabetes**<br>**- uncomplicated (ICD012)** | DE10, DE11, DE100, DE101-DE1019, DE109, DE110-DE1109, DE111-DE1119, DE119, DE120, DE121, DE129, DE130-DE1309, DE131-DE1319, DE139, DE140-DE1409, DE141 – DE1419, DE149 |
| **- complicated (ICD013)** | DE102-DE1029, DE103-DE1039, DE104-DE1049, DE105-DE1059,<br>DE106-DE1069, DE107-DE1079, DE108, DE112-DE1129, DE113- DE1139, DE114-DE1149, DE115-DE1159, DE116-DE1169, DE117-DE1179, DE118-DE1189, DE122-DE1229, DE123-DE1239, DE124-DE1249, DE125-DE1259, DE126-DE1269, DE127-DE1279, DE128- DE1289, DE132-DE1329, DE133-DE1339, DE134-DE1349, DE135-DE1359, DE136-DE1369,<br>DE137-DE1379, DE138, DE142-DE1429, DE143-DE1439, DE144-DE1449, DE145-DE1459, DE146-DE1469, DE147-DE1479, DE148 |
| **Major psychiatric disorder (ICD037)** | DF20-DF2099, DF22-DF229, DF25-DF2599, DF28-DF289, DF29-DF299, DF302-DF3029, DF31- DF3199, DF321-DF3219, DF322-DF3229, DF323-DF3239, DF328-DF3289, DF329-DF3299, DF33-DF3399, DF39-DF399 |

*M3-index, Measuring Mulitmorbidity index
†ICD10, Internation Classification of Disease 10th revision

| **Table S4** Drug groups and ATC*-codes | |
|---|---|
| **Drug category** | **ATC* code(s)** |
| **Systemic antibiotics** | **J01** (antibacterials for systemic use)<br>**J04** (antimycobacterials) |
| **NSAID** | **M01A** |
| **RAAS acting agents + potassium sparing diuretics** | **C09** (renin-angiotensin acting agents, including ACE-inhibitors, angiotensin receptor blockers and renin-inhibitors)<br>**C03D** (aldosterone antagonists and other potassium-sparing diuretics)<br>**C03E** (Diuretics and potassium sparing agents in combination) |
| **Beta-blocker or Cardiac glycoside** | **C07** (Beta blocker)<br>**C01A** (Cardiac glycoside) |
| **Potassium supplement** | **A12BA** |
| **Corticosteroids (mineralo- and glucocorticoids)** | **H02A** (plain)<br>**H02B** (combination) |

*ATC, Anatomical Therapeutic Classification

**<u>Table S5</u>** Missing data. Number of patient contacts with missing registration of the different acute parameters in patient groups defined by potassium measurement at emergency department arrival. The data is presented as frequencies and percentages of total contacts in each group.

| | Total | [K+] 3.5–4.4 mM | [K+] 4.5–4.9 mM | [K+] 5.0–5.9 mM | [K+] ≥6.0 mM |
|---|---|---|---|---|---|
| **Heart rate** | 129,412 (23.1%) | 115,103 (23.0%) | 11,106 (24.1%) | 2,773 (23.1%) | 430 (21.2%) |
| **Systolic Blood pressure** | 130,929 (23.3%) | 116,394 (23.2%) | 11,253 (24.4%) | 2,827 (23.5%) | 455 (22.5%) |
| **Oxygen Saturation** | 129,759 (23.1%) | 115,413 (23.0%) | 11,123 (24.1%) | 2,789 (23.2%) | 434 (21.4%) |
| **Body Temperature** | 139,271 (24.8%) | 123,740 (24.7%) | 11,905 (25.8%) | 3,088 (25.7%) | 538 (26.6%) |
| **Respiratory rate** | 134,067 (23.9%) | 119,090 (23.8%) | 11,531 (25.0%) | 2,932 (24.4%) | 514 (25.4%) |
| **p-[Creatinine]** | 4,180 (0.7%) | 3,630 (0.7%) | 371 (0.8%) | 137 (1.1%) | 42 (2.1%) |
| **p-[Na+]** | 155 (0.03%) | 131 (0.03%) | 16 (0.03%) | <10 (<0.05%) | <10 (<0.05%) |
